# Performance of the Illumina Infinium MethylationEPIC v.2 array with low DNA input from Swedish neonatal dry blood spots

**DOI:** 10.1101/2025.03.10.25323486

**Authors:** Gustaf Brander, Håkan Karlsson, Christina Dalman, Jonas Bybjerg-Grauholm, James J Crowley, David Mataix-Cols

## Abstract

The Swedish Phenylketonuria (PKU) screening biobank contains neonatal blood spots from over five million individuals born in Sweden since 1975, offering a unique research resource. While its value has been proven in several research areas, such as genetics, the feasibility of DNA methylation studies using archived dried blood spots remains uncertain. Here, we selected samples from seven random individuals born between 1985 and 2003 to evaluate the performance of the Illumina Infinium MethylationEPIC v2.0 array on DNA extracted from PKU blood spots. Despite DNA input quantities as low as 19.2 ng, probe call rates remained exceptionally high (mean 99.76% at p<0.01), with robust methylation site coverage across all samples. These findings challenge conventional assumptions about the DNA quantity and quality required for methylation arrays and demonstrate that decades-old neonatal blood spots can yield high-quality epigenetic data.

---

The Swedish Phenylketonuria (PKU) screening biobank is a unique and comprehensive resource containing neonatal blood spots from all children born in Sweden since 1975.^1^ With approximately 115,000 newborns screened annually, this biobank has accumulated samples from over five million individuals. Each sample consists of four dried blood spots on filter paper (Guthrie cards) collected within the first few days of life. It is accompanied by detailed metadata, including maternal information, birth details, and precise collection timing.

This systematic collection, initiated for screening over twenty congenital disorders but preserved under the Swedish Biobank Law of 2003 for medical care and ethically approved research, has already demonstrated its utility in genetic studies. For instance, the PKU neonatal blood spots have been used to successfully perform genotyping,^2^ exome sequencing,^3^ metabolics profiling,^4^ and protein analysis studies.^5^ Another study revealed only modest RNA decay over a 20-year period.^6^ A few studies have shown that the quality of DNA methylation data from dried blood spot samples were comparable to the quality from fresh blood samples,^7–9^ with Hollegaard et al. (2013) notably using samples stored for over two decades.

Whether the Swedish PKU blood spots are also suitable for methylation analyses is unclear. The Illumina Infinium MethylationEPIC v2.0 array requires 250 ng of DNA^10^, but a comprehensive evaluation of the chip showed high correlations between data from 250 ng of DNA and that from low DNA input.^11^ This suggests that the chip should yield meaningful results with much smaller amounts of DNA. Recent studies have confirmed the feasibility of methylation arrays with low DNA input using optimal sample types. Notably, a study comparing different DNA input tissues to detect oncogenic methylation achieved reliable results with 50 ng DNA from formalin fixed samples and 10 ng DNA from fresh blood samples using the Illumina Infinium HD assay.^12^

Here, we aimed to evaluate the feasibility of using the Illumina Infinium MethylationEPIC v.2 assay under far more challenging conditions. We focused on samples collected since 1981, when the Swedish PKU biobank started storing blood spots in a climate-controlled environment of 4°C and 30% humidity. Prior to 1981, samples were stored at room temperature, which may not be ideal conditions for methylation studies. The study was approved by the Swedish Ethical Review Authority (dnr: 2016-1130-31-2; amendment dnr: 2024-08257-02) without requiring informed consent from individual participants.

We randomly selected PKU blood spots from 7 individuals born across a span of 18 years (1985 to 2003). **Figure 1** shows the blood spot processing procedure. First, two 3 mm punches from each blood spot were punched into 96-well plates. Second, proteins and metabolites were eluted by incubating punches in 200 μl 1x phosphate buffered saline for 2 hours at room temperature on a rotary shaker (900 rpm). This eluate contains proteins such as acute phase proteins and antibodies and other metabolites (e.g., vitamin D, cytokines), which are banked for future studies. Third, DNA was extracted using the Highprep Blood & Tissue DNA kit from MagBio Genomics. The extraction was performed as an automated process using the BioMek I7 Automated Liquid Handler from Beckman Coulter Life Sciences. This yielded ∼100-200 ng of DNA per sample, 15-30 ng of which was used for genotyping. The remaining DNA was used for epigenetic profiling.

**Figure 1.**
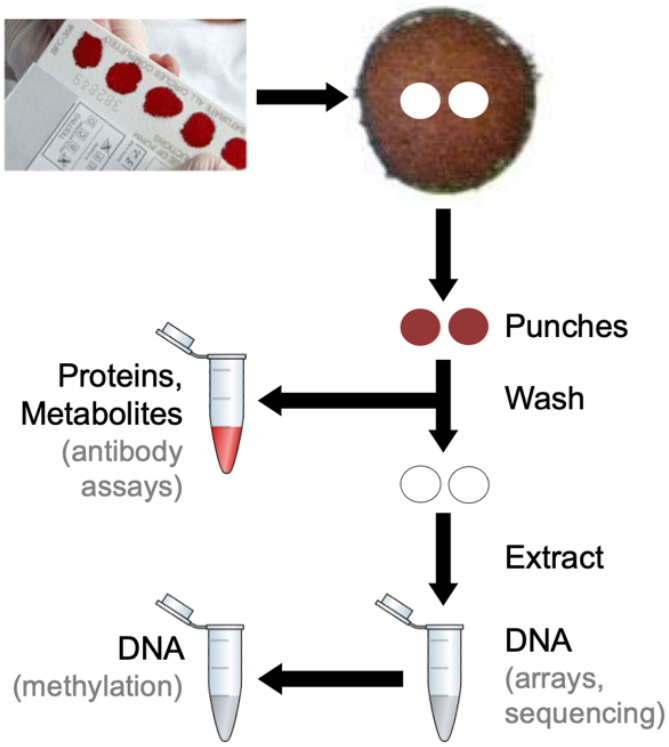
Blood spot processing procedure.

Bisulfite conversion, using the EZ-96 DNA Methylation-Lightning Kit (D5033), and methylation analysis, quantifying ∼930,000 CpG sites using The Illumina Infinium MethylationEPIC v.2 array (Illumina, San Diego, USA), were conducted at the National Genomic Infrastructure (NGI; https://www.scilifelab.se/units/ngi/) in Uppsala, Sweden.

The results are summarized in **Table 1**. DNA input quantities ranged from as low as 19.2 ng to 99.2 ng. Despite the age and storage conditions of these samples, our probe call rates at detection level p<0.01 ranged from 99.42% to 99.89%, with even higher rates (99.72% to 99.93%) at p<0.05. We detected between 931,598 and 935,979 methylation sites at p<0.01, demonstrating robust coverage across all samples regardless of DNA input quantity or sample age.

**Table 1.** Number of DNA methylation sites for each sample of the pilot, at detection levels p<0.01 and p<0.05. The value of the detection p value for each marker loci per sample is calculated by dividing the number of marker loci with detection p value <0.01 by the total of 937,055 marker loci on the Illumina Methylation EPIC array. All samples included for analysis in this pilot passed this limit.

| ID | Detected sites (0.01) | Probe Call Rate (0.01) | Detected sites (0.05) | Probe Call Rate (0.05) | Estimated DNA input (ng) |
| --- | --- | --- | --- | --- | --- |
| 1 | 935,314 | 0.9981 | 936,031 | 0.9989 | 51.0 |
| 2 | 931,598 | 0.9942 | 934,449 | 0.9972 | 19.2 |
| 3 | 934,882 | 0.9977 | 935,844 | 0.9987 | 35.2 |
| 4 | 935,979 | 0.9989 | 936,429 | 0.9993 | 70.4 |
| 5 | 935,549 | 0.9984 | 936,175 | 0.9991 | 32.0 |
| 6 | 935,372 | 0.9982 | 935,884 | 0.9988 | 99.2 |
| 7 | 934,661 | 0.9974 | 935,748 | 0.9986 | 48.0 |

These excellent results challenge conventional assumptions about the minimum quantity and quality requirements for methylation analyses using archived dried blood spots. This creates opportunities for large-scale longitudinal research using material from the Swedish PKU biobank while preserving precious sample material for future analyses. A remaining question for the future is whether blood spots stored at room temperature before 1981 are also suitable for methylation analyses.

## Data availability statement

The data cannot be shared due to gdpr and Swedish legislation.

## Ackhowledgements

*Methylation analysis by genotyping was performed by the SNP&SEQ Technology Platform in Uppsala. The facility is part of the National Genomics Infrastructure (NGI) Sweden and Science for Life Laboratory. The SNP&SEQ Technology Platform is also supported by the Swedish Research Council and the Knut and Alice Wallenberg Foundation*.

## Discolsures

Prof Mataix-Cols receives royalties for contributing articles to UpToDate, Inc. and is part-owner of Scandinavian E-Health AB, all outside the current work.

